# COVID-19 pediatric mortality rates are heterogeneous between countries

**DOI:** 10.1101/2020.09.17.20196832

**Authors:** Nadia González-García, América Liliana Miranda-Lora, Juan Garduño-Espinosa, Javier T. Granados-Riverón, Jorge Fernando Méndez-Galván, Jaime Nieto-Zermeño, María F Castilla-Peón

**Author notes:** Autor de correspondencia: María Fernanda Castilla Peón. Dirección de Investigación. Hospital Infantil de México Federico Gómez. Dirección: Dr. Márquez No. 162. Colonia Doctores, Delegación Cuauhtémoc, CP 06720, Ciudad de México. Teléfono 5228-9917. Correo electrónico.

## Abstract

**Introduction:** Severe COVID-19 is infrequent in children, with a lethality rate of about 0.08%. This study aims to explore differences in the pediatric mortality rate between countries.

**Methods:** Countries with populations over 5 million that report COVID-19 deaths disaggregated data by quinquennial or decennial age groups were analyzed. Data were extracted from COVID-19 Cases and Deaths by Age Database, national ministries of health, and the World Health Organization.

**Results:** 23 countries were included in the analysis. Pediatric mortality varied from 0 to 12.1 deaths per million people of the corresponding age group, with the highest rate in Peru. In most countries, deaths were more frequent in the 0-4 years old age group, except for Brazil. The pediatric/ general COVID-19 mortality showed a great variation between countries and ranged from 0 (Republic of Korea) to 10.4% (India). Pediatric and Pediatric/general COVID mortality have a strong correlation with 2018 neonatal mortality (r=0.77, p<0.001 and r= 0.88, p<0.001 respectively), while it has a moderate or absent (r=0.47, p=0.02 and r=0.19, p=0.38, respectively) correlation with COVID-19 mortality in the general population.

**Conclusions:** There is an important heterogenicity in pediatric COVI-19 mortality between countries that parallels historical neonatal mortality. Neonatal mortality is a known index of the quality of a country’s Health System which points to the importance of social determinants of health in pediatric COVID-19 mortality disparities, an issue which should be further explored.

## Introduction

Since the first cases of coronavirus disease, 2019 (COVID-19) appeared in Wuhan, China, at the end of 2019, morbidity and mortality by SARS-CoV-2 have been significant in countries within every continent. Mortality is mostly concentrated in advanced age groups and the adult population with ongoing comorbidities (1-5). Children and adolescents constitute 2% and 9.5% of all reported cases in Europe and the USA, respectively. Severe COVID-19 is infrequent in children and intensive care unit admissions regarding this age group have been reported to be about 2% and lethality rate about 0.08% (6). Besides, preliminary evidence suggests that both ethnicity (Black and Hispanic) and age (under 1 month and early adolescence [10-14 years]) are associated with admission to a critical care unit. Moreover, children from low-income families or non-white ethnicity are more likely to test positive for SARS-CoV-2 than those of white ethnicity and high-income (7, 8).

To date, COVID-19 epidemiologic data in the pediatric population has been published by some countries but there is a scarcity of analysis comparing different populations. The COVID-19 mortality rate in the population under 18 years of age, might be heterogeneous between countries with different incomes rates and ethnicities. Our aim with this brief report is to explore the differences in the COVID-19 pediatric mortality rate between countries.

## Methods

We included information about countries with populations over 5 million that reported COVID-19 deaths, disaggregated by quinquennial or decennial age groups. Data about confirmed COVID-19 death counts were consulted in the COVID-19 Cases and Deaths by Age Database (COVerAge-DB) (9), COVerAge-DB collects age- and sex-specific cumulative cases, deaths, and tests from official reports from multiple countries worldwide and for several subpopulations. Data from Argentina, Canada, and Peru, and Mexico were consulted from the local Ministries of Health (10-13) Data from countries that disaggregate data by decennial age groups, were estimated according to the countries age structure available data.

For the computation of age-specific mortality rates, we obtained both the total and quinquennial age groups estimated populations for 2018 or the latest available year from the United Nations Statistics Division Web site, (14). Data for general COVID-19 mortality were extracted from the World Health Organization (WHO) reports (15).

In an exploratory analysis, we also calculated the Spearman’s correlation coefficient between pediatric COVID-19 mortality rates with general mortality rates and newborn mortality by any cause in 2018 (before the COVID-19 pandemic). STATA 13.0 ® was used for the analysis.

## Results

We included 23 countries in the analysis. Table 1 shows the general mortality and pediatric mortality rates from COVID-19. Specifically, 63% of deaths in the population of <15 years of age at the time of the study occurred in India (n = 1,622). However, when adjusting the mortality per million people for this age group, the highest rates were observed in Latin American countries (Peru, Brazil, Ecuador, and Mexico). Although Peru also has the highest overall mortality rate from COVID-19, it is followed by European countries (United Kingdom, Spain, and Italy). On the other hand, the highest pediatric to general population ratio of COVID-19 mortality was identified in Asian countries (India, Indonesia, and the Philippines).

**Table 1.**
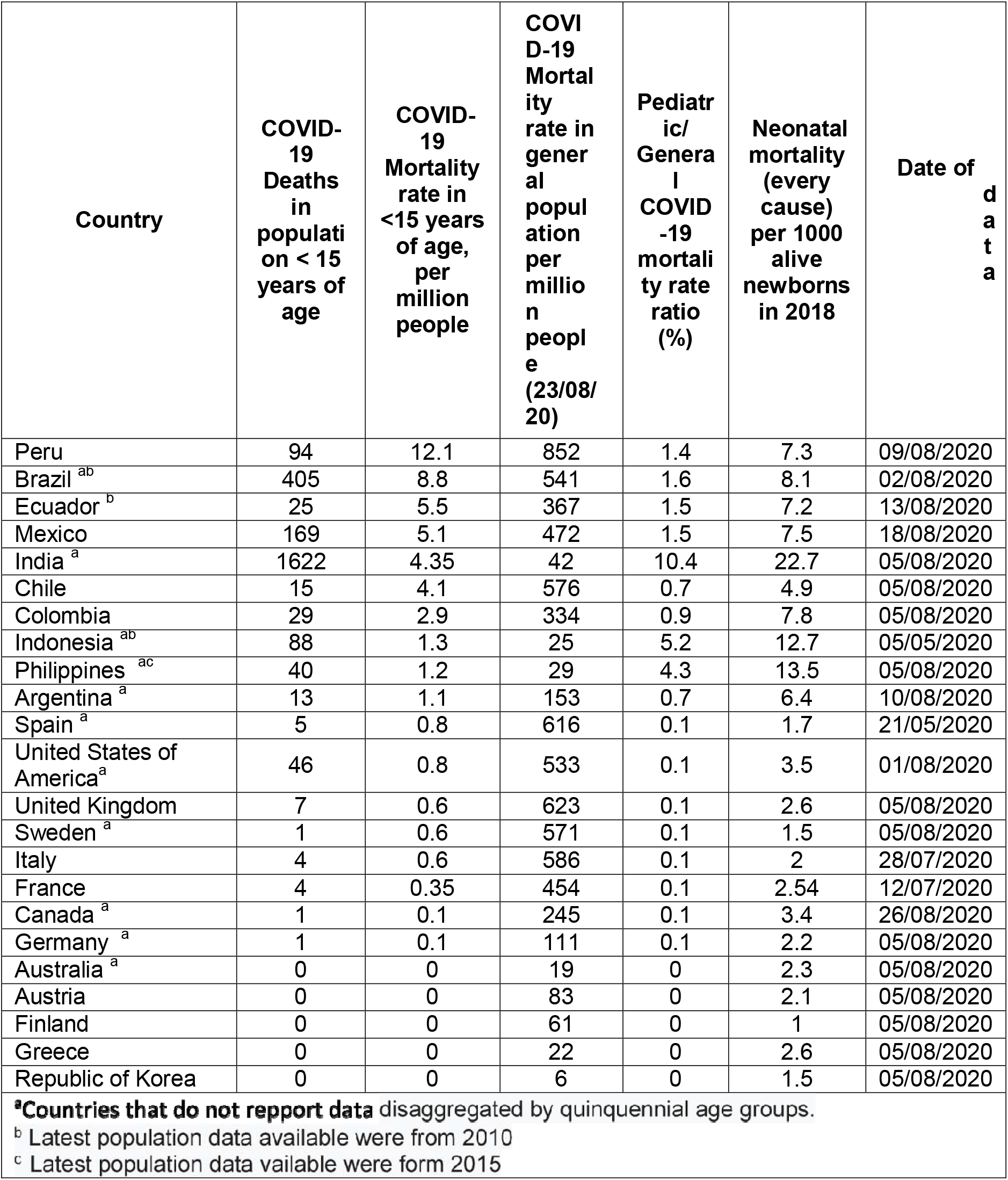
Mortality indices by COVID-19 and neonatal mortality in 2018 by country.

| Country | COVID-19 Deaths in population < 15 years of age | COVID-19 Mortality rate in <15 years of age, per million people | COVID-19 Mortality rate in general population per million people (23/08/20) | Pediatric/General COVID-19 mortality rate ratio (%) | Neonatal mortality (every cause) per 1000 alive newborns in 2018 | Date of data |
| --- | --- | --- | --- | --- | --- | --- |
| Peru | 94 | 12.1 | 852 | 1.4 | 7.3 | 09/08/2020 |
| Brazil <sup>ab</sup> | 405 | 8.8 | 541 | 1.6 | 8.1 | 02/08/2020 |
| Ecuador <sup>b</sup> | 25 | 5.5 | 367 | 1.5 | 7.2 | 13/08/2020 |
| Mexico | 169 | 5.1 | 472 | 1.5 | 7.5 | 18/08/2020 |
| India <sup>a</sup> | 1622 | 4.35 | 42 | 10.4 | 22.7 | 05/08/2020 |
| Chile | 15 | 4.1 | 576 | 0.7 | 4.9 | 05/08/2020 |
| Colombia | 29 | 2.9 | 334 | 0.9 | 7.8 | 05/08/2020 |
| Indonesia <sup>ab</sup> | 88 | 1.3 | 25 | 5.2 | 12.7 | 05/05/2020 |
| Philippines <sup>ac</sup> | 40 | 1.2 | 29 | 4.3 | 13.5 | 05/08/2020 |
| Argentina <sup>a</sup> | 13 | 1.1 | 153 | 0.7 | 6.4 | 10/08/2020 |
| Spain <sup>a</sup> | 5 | 0.8 | 616 | 0.1 | 1.7 | 21/05/2020 |
| United States of America<sup>a</sup> | 46 | 0.8 | 533 | 0.1 | 3.5 | 01/08/2020 |
| United Kingdom | 7 | 0.6 | 623 | 0.1 | 2.6 | 05/08/2020 |
| Sweden <sup>a</sup> | 1 | 0.6 | 571 | 0.1 | 1.5 | 05/08/2020 |
| Italy | 4 | 0.6 | 586 | 0.1 | 2 | 28/07/2020 |
| France | 4 | 0.35 | 454 | 0.1 | 2.54 | 12/07/2020 |
| Canada <sup>a</sup> | 1 | 0.1 | 245 | 0.1 | 3.4 | 26/08/2020 |
| Germany <sup>a</sup> | 1 | 0.1 | 111 | 0.1 | 2.2 | 05/08/2020 |
| Australia <sup>a</sup> | 0 | 0 | 19 | 0 | 2.3 | 05/08/2020 |
| Austria | 0 | 0 | 83 | 0 | 2.1 | 05/08/2020 |
| Finland | 0 | 0 | 61 | 0 | 1 | 05/08/2020 |
| Greece | 0 | 0 | 22 | 0 | 2.6 | 05/08/2020 |
| Republic of Korea | 0 | 0 | 6 | 0 | 1.5 | 05/08/2020 |

<sup>a</sup>Countries that do not report data disaggregated by quinquennial age groups.
<sup>b</sup> Latest population data available were from 2010
<sup>c</sup> Latest population data available were form 2015

Table 2 shows the pediatric mortality by quinquennial age groups. The highest mortality in children under 10 years old was observed in Peru and the highest mortality in children older than 10 years old was in Brazil.

**Table 2.** COVID-19 Mortality rate by quinquennial age groups and country.

| *Country* | **COVID-19 Deaths in the population aged 0-4 years** | **COVID-19 Mortality rate in the population aged 0-4 years (per million)** | **COVID-19 Deaths in the population aged 5-9 years (per million)** | **COVID-19 Mortality rate in the population aged 5-9 years (per million)** | **COVID-19 Deaths in the population aged 10-14 years** | **COVID-19 Mortality rate in the population aged 10-14 years (per million)** |
| --- | --- | --- | --- | --- | --- | --- |
| Peru | 40 | 16.04 | 29 | 10.96 | 25 | 9.57 |
| India <sup>a</sup> | 1519.1 | 13.47 | 20.5 | 0.16 | 82.2 | 0.62 |
| Mexico | 108 | 9.85 | 25 | 2.25 | 36 | 3.22 |
| Ecuador <sup>b</sup> | 11 | 7.52 | 7 | 4.58 | 7 | 4.55 |
| Chile | 9 | 7.24 | 3 | 2.40 | 3 | 2.52 |
| Colombia | 15 | 4.94 | 9 | 2.70 | 5 | 1.38 |
| Brazil <sup>ab</sup> | 38.2 | 2.77 | 103.8 | 6.93 | 263.1 | 15.33 |
| Philippines <sup>ac</sup> | 26 | 2.40 | 7 | 0.65 | 7 | 0.67 |
| Indonesia <sup>ab</sup> | 47.4 | 2.09 | 22.3 | 0.96 | 18.4 | 0.81 |
| United States of America <sup>a</sup> | 25.3 | 1.28 | 8.1 | 0.40 | 12.2 | 0.58 |
| Argentina (5) <sup>a</sup> | 3.5 | 0.94 | 3.5 | 0.94 | 5.5 | 1.56 |
| Italy | 2 | 0.82 | 2 | 0.82 | 0 | 0 |
| United Kingdom | 3 | 0.76 | 1 | 0.24 | 3 | 0.79 |
| Sweden <sup>a</sup> | 0.4 | 0.66 | 0.6 | 0.98 | 0 | 0 |
| Spain <sup>a</sup> | 0.9 | 0.43 | 1.9 | 0.78 | 2.5 | 1.02 |
| France | 1 | 0.28 | 2 | 0.5 | 1 | 0.25 |
| Germany <sup>a</sup> | 1 | 0.26 | 0 | 0.00 | 0 | 0 |
| Canada (6) <sup>a</sup> | 0.25 | 0.13 | 0.25 | 0.12 | 0.25 | 0.13 |
| Australia | 0 | 0 | 0 | 0 | 0 | 0 |
| Austria | 0 | 0 | 0 | 0 | 0 | 0 |
| Finland | 0 | 0 | 0 |  | 0 | 0 |
| Greece | 0 | 0 | 0 | 0 | 0 | 0 |
| Republic of Korea | 0 | 0 | 0 | 0 | 0 | 0 |

<sup>a</sup>Countries that do not report data disaggregated by quinquennial age groups.
<sup>b</sup> Latest population data available were from 2010
<sup>c</sup> Latest population data available were from 2015

We found a significant strong correlation between both the COVID-19 pediatric mortality and the pediatric/general COVID-19 mortality ratio and the 2018 neonatal mortality (r=0.77, p<0.001 and r= 0.88, p<0.001 respectively), while there was a moderate or absent correlation with COVID-19 mortality in the general population. (r=0.47, p=0.02 and r=0.19, p=0.38, respectively) (Table 3).

**Table 3.**
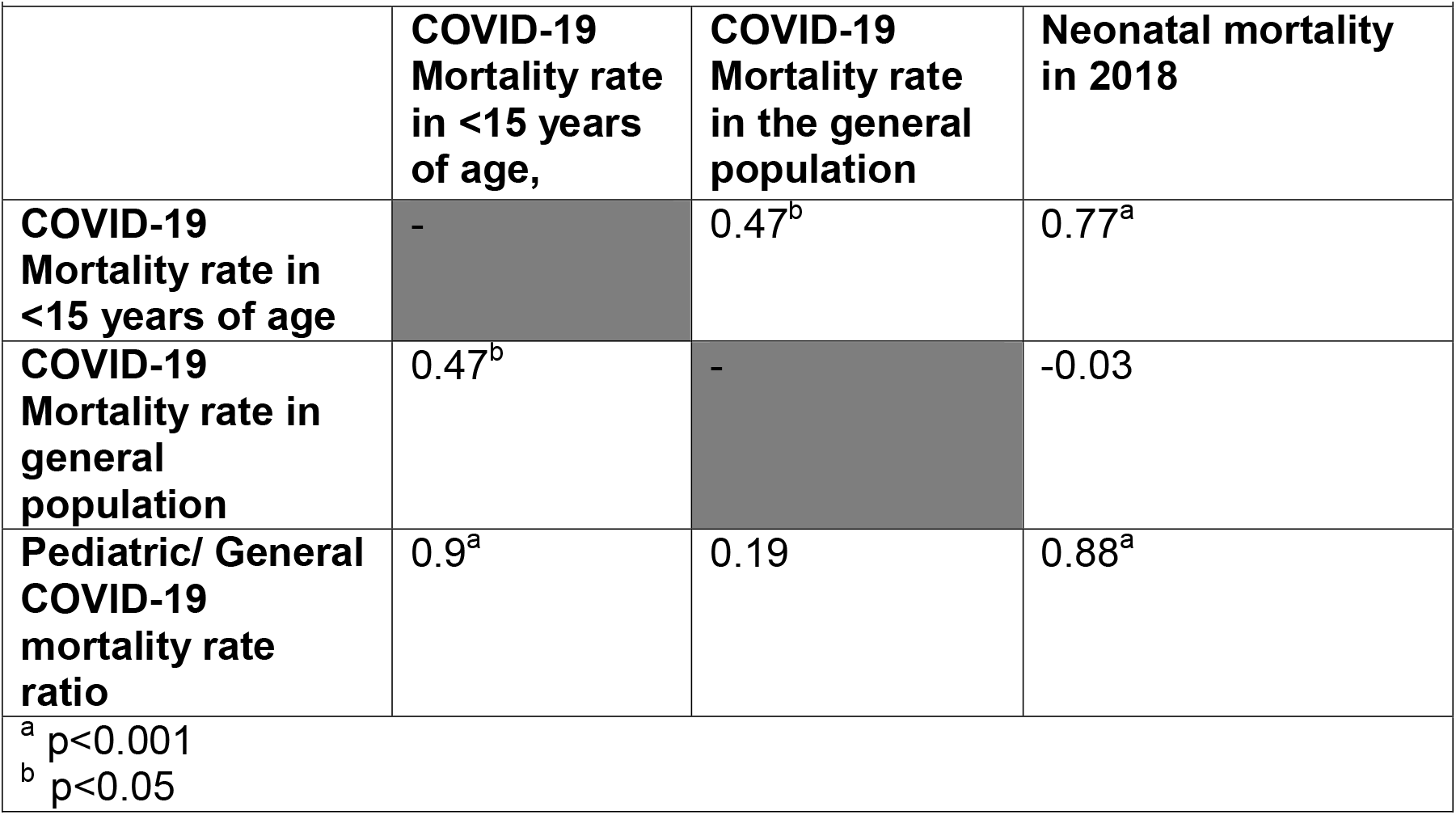
Spearman correlation coefficient between pediatric COVID-19 mortality, general COVID-19 mortality, Neonatal mortality in 2018, and Pediatric/General COVID-19 mortality rate ratio.

## Discussion

Despite COVID-19 mortality in children is minimal in comparison with that in the adult population, there is significant heterogeneity between countries. Several factors should be explored to explain this variability. This report was elaborated with available data from different sources and differences in reporting systems of epidemiological information may be accountable for some of the variation.

Remarkably, the highest pediatric mortality rates are among countries of upper-middle-income countries in contrast with high-income countries (data for lower and lower-middle-income countries were not available). This is true even for high-income countries that have suffered from high mortality rates in the general population which, despite this, have a low pediatric/general COVID mortality rate ratio (adjusted for age structure).

In most countries, the COVID mortality rate in the pediatric population is concentrated in the under 5-year-old population. Brazil has a disproportionately high mortality rate in adolescents and the causes of this must be studied. In Mexico, about half of the deaths in the 0-4 age group are in infants < 1-year-old. It is important to have disaggregated data for age to estimate the share of child mortality which corresponds to neonatal and infant mortality.

Child, infant, and neonatal mortality are known indicators of the quality of health care systems(16). It is noticeable that pediatric COVID mortality and pediatric/general COVID-19 rate ratio are highly correlated with historical basal neonatal mortality while it is only moderately correlated with general COVID-19 mortality. These findings suggest an important role of social determinants of health as well as the quality of health care systems in discrepancies of pediatric COVID-19 mortality rates between countries. The relative importance of this set of factors over biological factors remains to be established.

## Data Availability

Data are available in the websites cited in the manuscript

https://osf.io/mpwjq/

https://www.argentina.gob.ar/salud/coronavirus-COVID-19/sala-situacion

https://www.datosabiertos.gob.pe/dataset/fallecidos-por-covid-19-ministerio-de-salud-minsa

https://www.gob.mx/salud/documentos/datos-abiertos-152127

https://covid19.who.int/

